# Empowering digital health management with on-device large language models for glucose prediction

**DOI:** 10.1101/2025.07.12.25331188

**Authors:** Taiyu Zhu, Joanna Howson, Alejo Nevado-Holgado

## Abstract

Long-term management of chronic diseases such as diabetes is increasingly based on wearable technologies, particularly continuous glucose monitoring (CGM), integrated with smartphone-based digital health systems. When combined with artificial intelligence, especially deep learning, these systems offer highly personalized decision support, including glucose prediction. Although large language models (LLMs) have demonstrated strong performance across various healthcare tasks, their integration into day-to-day digital health remains limited, primarily due to privacy concerns associated with transmitting sensitive data to remote servers. Recent advances in lightweight LLMs create new opportunities for secure and local deployment. In this study, we first evaluated the zero-shot glucose prediction performance of eight pretrained lightweight LLMs across multiple model families. None achieved clinically viable outputs, highlighting the need for domain-specific adaptation. To address this, we propose GluLLM, a multimodal adapter-based framework that enhances pretrained LLMs for on-device glucose forecasting. GluLLM integrates CGM data, daily activity logs, and electronic health records using customized encoder and decoder modules while preserving the foundational capabilities of pretrained LLMs. We trained and evaluated GluLLM on the REPLACE-BG dataset, which includes 226 individuals with type 1 diabetes, and validated it on an external cohort comprising 207 individuals with type 2 diabetes or without diabetes. Compared with 15 state-of-the-art deep learning baseline models for time series prediction, GluLLM with a LLaMA 3.2 1B backbone achieved superior prediction performance, demonstrating the lowest root mean square error and the highest sensitivity for hypoglycemia detection in both datasets. Furthermore, the deployment of GluLLM on two smartphone platforms demonstrated feasible computational requirements with acceptable CPU and memory usage, highlighting its practical utility for real-world LLM-driven digital health management.

## Introduction

Characterized by insufficient insulin production or dysfunctional beta cells in the pancreas, diabetes remains one of the most significant global health challenges [1]. It is estimated that over half a billion people worldwide are living with diabetes [2], with approximately 90% diagnosed with type 2 diabetes (T2D) and around 10% with type 1 diabetes (T1D). Effective glucose regulation, or glycemic control, poses a substantial daily burden for individuals with T1D or insulin-dependent T2D, which requires frequent glucose monitoring and multiple insulin interventions. Failure to maintain glucose levels within a therapeutic range can lead to a range of complications [3], such as cardiovascular disease, neuropathy, and retinopathy. Therefore, consistent glucose monitoring and precise interventions are essential for effective diabetes management. Significant advances in wearable and mobile technologies in the past decade have facilitated the widespread adoption of continuous glucose monitoring (CGM) systems [4]. CGM measures interstitial glucose levels, which represent the glucose present in the fluid between cells, using a minimally invasive sensor inserted under the skin, typically on the abdomen or upper arm. These readings are then converted into real-time estimates of blood glucose levels, generally updated every five minutes, providing users with a dynamic and continuous view of glycemic trends. Multiple large-scale clinical studies have demonstrated that CGM technology substantially improves patient outcomes, reduces the burden of daily management, and improves quality of life through more comprehensive glycemic monitoring and reduced measurement invasiveness in both T1D and T2D [5–8]. Despite advances in diabetes management technologies, the effectiveness of real-world interventions remains limited by several physiological and temporal constraints. There is a notable time delay of five to ten minutes between changes in blood glucose levels and their detection by interstitial CGM sensors [9]. Insulin therapy introduces additional delays due to its pharmacokinetics, as most formulations begin acting about 15 minutes after administration and may continue to exert effects for several hours [10]. Dietary glucose absorption also varies significantly across individuals and food types, adding further complexity. One particularly important challenge is the detection of hypoglycemia, which often occurs without noticeable symptoms and can escalate to life-threatening episodes, such as seizures, loss of consciousness, or stroke [11]. These combined temporal factors emphasize the importance of proactive diabetes management strategies, with accurate glucose prediction serving as a crucial foundation for timely and effective intervention.

However, achieving reliable glucose prediction is inherently difficult due to substantial inter- and intra-individual variability [12, 13]. Various daily activities, such as food intake, insulin administration, and physical exercise, affect glucose levels, while individual characteristics, such as HbA1c and body mass index (BMI), are also closely associated with glucose dynamics [14]. Recent advances in artificial intelligence (AI), particularly deep learning [15], have enabled the effective use of CGM time series by leveraging their complex and heterogeneous nature. In the literature, a variety of deep neural network architectures designed for sequence modeling have been proposed to address this task, including convolutional neural networks (CNNs) [16], recurrent neural networks (RNNs) such as long short-term memory (LSTM) [17] and gated recurrent unit (GRU) [18–20] models, and more recently Transformer-based models [21]. These deep learning algorithms are capable of predicting glucose levels 30 to 60 minutes in advance, allowing individuals sufficient time to implement proactive interventions to mitigate adverse glycemic events, which have demonstrated superior predictive accuracy, as measured by metrics including the root mean square error (RMSE) [15]. Among these approaches, a critical consideration is the trade-off between personalized models tailored to individuals and population models applicable across diverse users. Although most of the literature focuses on personalized models to capture individual glycemic patterns [17, 19, 20, 22], these approaches face substantial barriers to clinical deployment, including time-intensive data collection processes and complex regulatory requirements. Population models present a favorable alternative for real-world applications; however, the substantial inter-subject variability poses notable challenges for model generalization. Furthermore, many studies have adopted single prediction horizon frameworks (i.e., sequence-to-one) [15, 17, 19, 20, 22], which limit their effectiveness in supporting decision-making processes [23].

To address these challenges simultaneously, recently proposed large language models (LLMs) offer promising new directions, particularly for integrating complex clinical knowledge [24, 25]. First, their generalization ability enables a trained model to provide accurate predictions for previously unseen individuals without requiring further fine-tuning. Second, their sequence-to-sequence architecture allows the model to predict a sequence of future glucose levels based on historical glucose measurements and relevant contextual information. The potential of LLMs for time series applications has been demonstrated across numerous recent studies [26–30]. These approaches leverage learned token transitions and language-based embeddings to encode both instructions and temporal data, consistently achieving state-of-the-art performance on diverse prediction benchmarks. Despite this success, the application of LLMs to glucose management remains relatively unexplored. Only two studies have ventured into this area: one evaluated TimeLLM [26], an existing LLM-based forecasting model, for personalized glucose prediction [31], while another employed GPT-4 to analyze CGM profiles [32]. However, a key challenge in the implementation of LLM in healthcare is to address the security and privacy concerns associated with processing sensitive data on remote platforms based on the Internet [25]. This challenge has sparked a promising trend toward smaller and more efficient LLMs, which is already becoming a reality for local deployment on personal devices. Apple Intelligence launched on-device LLMs on the latest iPhones in July 2024 providing API access to app developers in June 2025. This was followed by Meta’s release of LlaMA 3.2 edge AI models in September 2024. This evolution is perfectly in line with the practices of the CGM system, where smartphone apps already play a central role in facilitating data visualization and clinical decision support [33], and have demonstrated the ability to effectively run deep learning algorithms for glucose prediction [19]. Therefore, smartphones represent a natural choice to implement LLM-based clinical decision support in glucose management applications.

In this paper, we first evaluate the zero-shot learning performance of lightweight LLMs for glucose prediction. Subsequently, after identifying the current limitations in their performance, we propose GluLLM, a novel model-agnostic adapter framework that significantly enhances the glucose prediction capabilities of pretrained LLMs, as shown in Fig. 1. This framework leverages demographic information and temporal dependencies through a mixed embedding strategy for personalized prediction, which consistently outperforms 15 state-of-the-art deep learning-based time series prediction methods, including models based on Transformer, multilayer perceptrons (MLPs), CNNs, and RNNs. Finally, we implement GluLLM on two smartphone platforms and analyze CPU and memory consumption. This enables a new paradigm in smartphone-based LLM-driven digital health systems with strong potential for adaptation across a broad spectrum of chronic disease management applications.

**Fig. 1:**
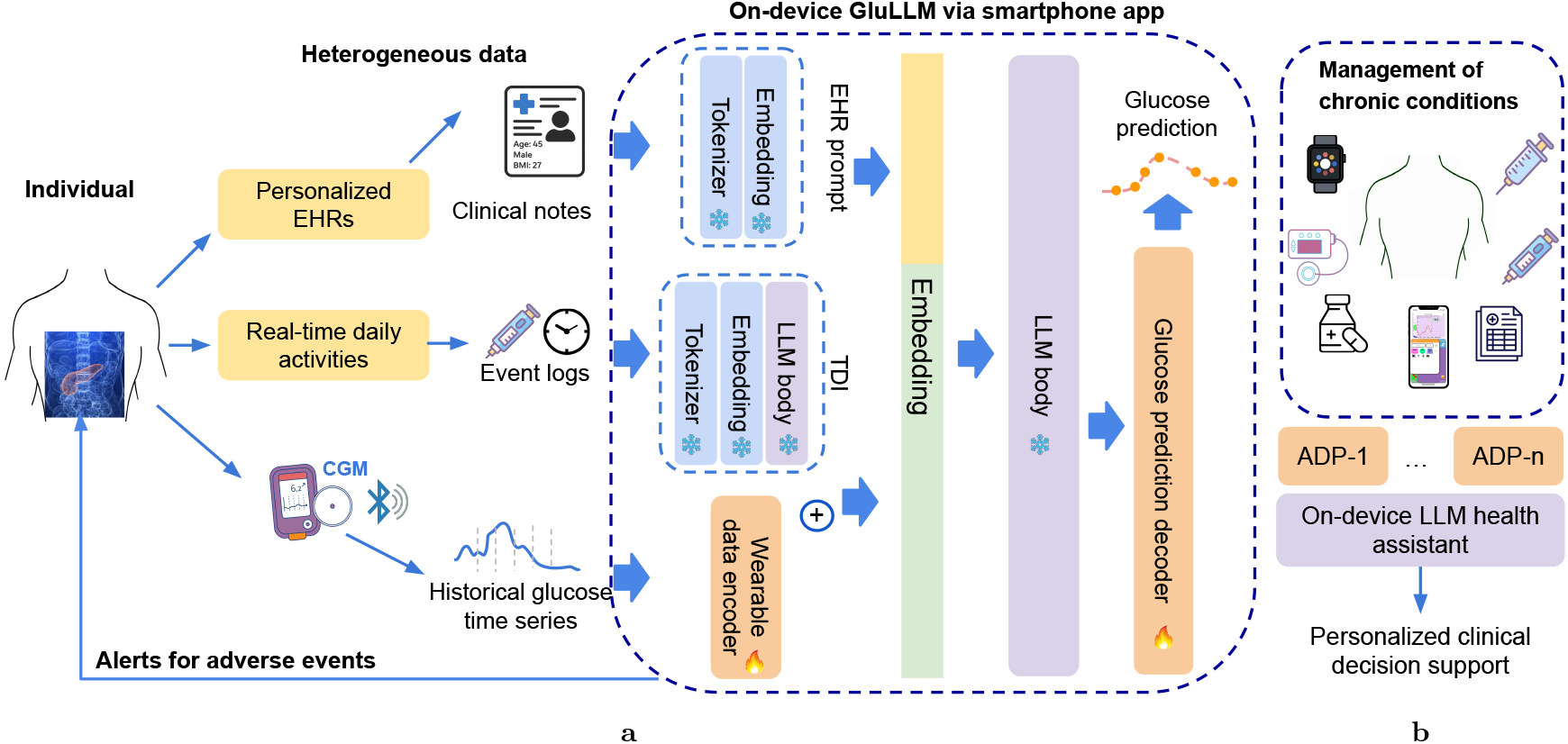
Integration of LLMs in chronic disease management through adaptive frameworks. **a**, Implementation of GluLLM on smartphone platforms that seamlessly integrate with digital health ecosystems for comprehensive glucose management. The system processes CGM data transmitted via Bluetooth, captures daily activities through event logs, and accesses electronic health records (EHRs) stored locally. All data streams are processed through specialized components of the GluLLM architecture, including EHR prompt, time dependent interpreter (TDI), and wearable data encoder, to deliver personalized insights and predictions. In the schematic, ice icons indicate preserved pretrained LLM elements that remain frozen, while fire icons denote adapter components requiring targeted training. **b** Extensibility of the framework through multiple specialized adapter (ADP), each equipped with dedicated encoder and decoder modules for handling diverse data types. This design enables management of multiple chronic conditions through a unified on-device LLM platform.

## Results

### Zero-short glucose prediction performance

Our initial experiment evaluated the zero-shot learning performance of LLMs for glucose prediction, which refers to the ability of a pretrained model to perform a specialized task without any additional fine-tuning or training. We constructed the following prompts that included comprehensive demographic information (age, sex, BMI, and HbA1c) alongside six hours of historical CGM readings. These structured prompts instructed the model to generate glucose predictions in a standardized format, allowing us to assess the baseline performance of pretrained LLMs.

“A [age]-year-old [sex] individual has an HbA1c of [HbA1c]% and a BMI of [BMI]. Based on their continuous glucose monitoring (CGM) data recorded every 5 minutes over the past 6 hours [historical_readings], predict their glucose levels for the next hour at 5-minute intervals.”

To ensure practical smartphone deployment of digital clinical decision support systems, we selected compact open-source models from leading LLM families, including LLaMA (Meta) [34], Mistral (Mistral AI) [35], Gemma (Google) [36], Phi (Microsoft) [37], OpenELM (Apple) [38], and GPT (OpenAI) [39]. Table 1 presents detailed information on model versions, parameter sizes, and response characteristics.

**Table 1:** Comparison of LLM performance on zero-shot glucose level prediction.

| Model | Size | Response quality |
| --- | --- | --- |
| LLaMA 3.2 | 1B | Attempted a response but misinterpreted the task, referring to blood pressure and concluding without a specific prediction. |
| LLaMA 3.2 | 3B | Initiated a structured statistical approach but contained calculation inaccuracies and did not produce a concrete prediction. |
| LLaMA 3.1 | 8B | Offered theoretical time series methodology discussion in multiple steps but avoided calculations, providing only a placeholder "0" as the final answer. |
| Mistral 0.2 | 7B | Started explaining analytical forecasting methods like linear regression in Python code but did not deliver any predictions. |
| Gemma 2 | 2B | Emphasized general medical considerations and disclaimers without directly addressing the prediction component of the task. |
| Phi-3.5 mini | 3.8B | Discussed machine learning strategies (e.g., RNNs with PyTorch) but the response lacked focus and did not complete the prediction. |
| OpenELM | 270M | Returned a series of other clinical features (e.g., blood pressure) but did not address the glucose prediction objective. |
| GPT-2 | 1.5B | Generated off-topic text discussing unrelated medical studies with no attempt at glucose prediction. |

Our evaluation revealed a significant performance gap: none of these LLMs demonstrated the capability to produce clinically viable glucose predictions. Rather than generating the requested numerical predictions, most models produced theoretical explanations of time series prediction methodologies (e.g., linear regression and RNNs) accompanied by Python implementations. These responses represent a substantial deviation from the required clinical functionality, requiring technical expertise beyond that possessed by most users and healthcare providers, and demonstrate a fundamental misalignment between the models’ generative tendencies and the specific requirements of prediction tasks. These findings highlight the critical limitations of applying general-purpose LLMs directly to specialized clinical tasks without appropriate adaptations.

### Enhancing glucose prediction in pretrained LLMs with GluLLM

To address the performance limitations of pretrained LLMs in glucose prediction, we propose the GluLLM framework (see Methods), which enhances predictive accuracy through targeted adaptation. Our results demonstrate that when augmented with GluLLM, all evaluated LLMs successfully provided viable glucose predictions with a one-hour forecast horizon. We conducted comprehensive performance comparisons between GluLLM and a diverse array of existing deep learning time series forecasting approaches. This comparison included ten state-of-the-art methods that, to our knowledge, are being applied to glucose prediction for the first time in this study. The baseline methods encompass three architectural categories: Transformer-based models including Autoformer [40], Crossformer [41], FEDformer [42], Informer [43], iTransformer [44], non-stationary Transformer (NST) [45], PatchTST [46], and vanilla Transformer [47]; MLP architectures such as N-Beats [48], DLinear [49], and TiDE [50]; convolutional neural networks including TCNs [16] and TimesNet [51]; and RNN architectures including LSTM [17] and GRU [19]. This extensive evaluation enables robust assessment of GluLLM’s performance relative to current methodological standards in time series forecasting.

To ensure a fair evaluation, all models underwent consistent training and assessment using the REPLACE-BG dataset [52]. We employed five-fold cross-validation for hyperparameter tuning and model training using a development set (*n* = 180), followed by performance evaluation on separate hold-out testing sets (*n* = 46). Dataset partitioning was conducted on an individual basis to prevent data leakage and mimic real-world scenarios with heterogeneous populations. To enhance the generalization capabilities of non-LLM models, we applied a model-agnostic metalearning framework for domain generalization [53], which has effectively mitigated the challenges of high inter-individual variability and improved glucose prediction performance as evidenced in our recent work [21]. Fig. 2 presents the global RMSE values comparing predicted sequences with actual CGM measurements over the next 60 minutes. The results demonstrate that LLaMA 3.2 models achieved superior performance, with the 1B version ranking at the top. Other LLMs in our evaluation exhibited performance comparable to the average level of deep learning baseline methods. Among the tested LLMs, OpenELM delivered the lowest performance, which may be attributed to its significantly smaller parameter count compared to other architectures.

**Fig. 2:**
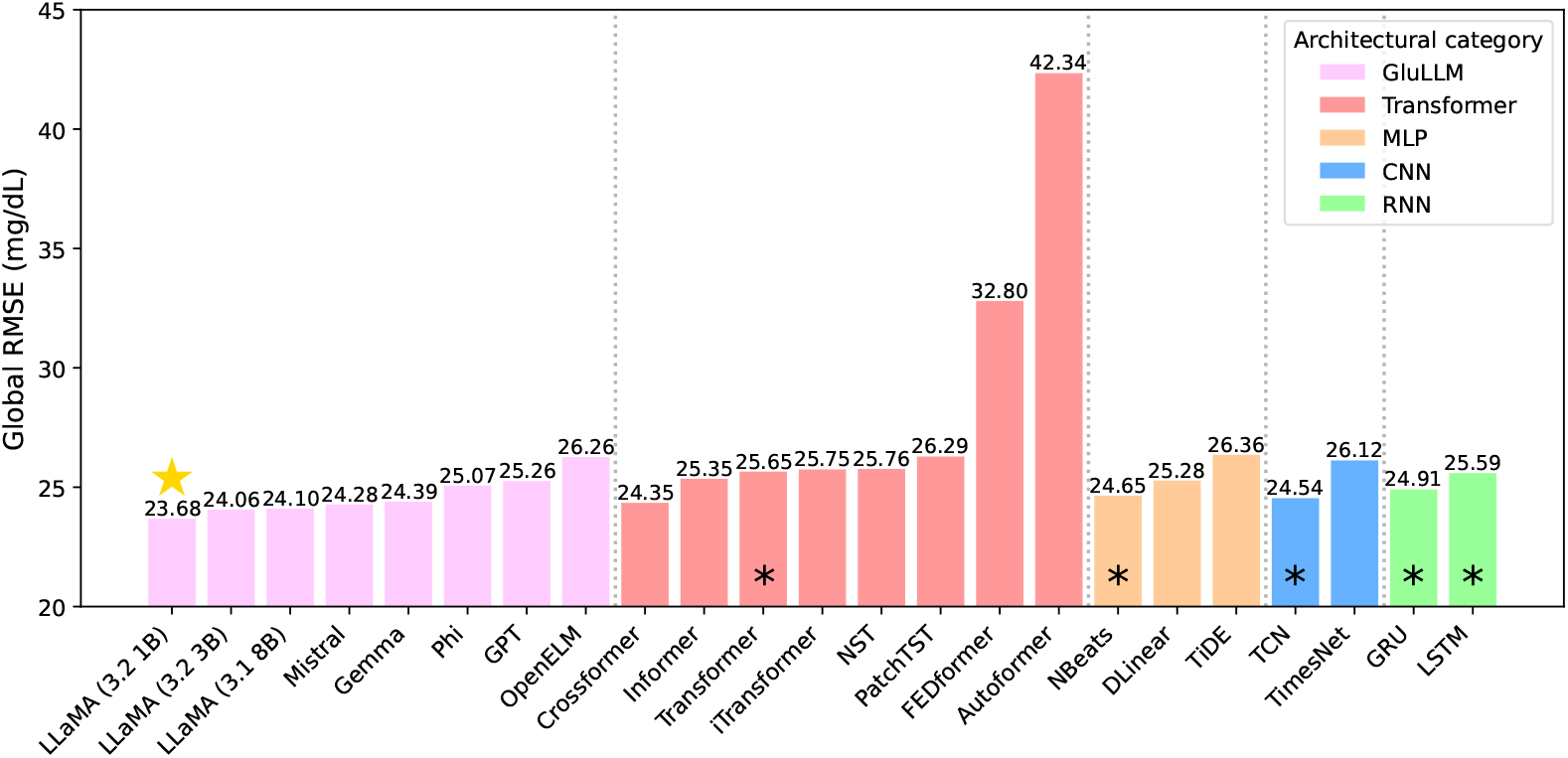
Global RMSE for glucose time series across different model architectures with a 60-minute prediction horizon. The color coding represents distinct architectural categories: violet for LLM-based models, light red for Transformer-based models, light yellow for MLP-based models, blue for CNN-based models, and green for RNN-based architectures. Black asterisk markers indicate models previously explored in glucose prediction literature, while the gold star highlights the best-performing model in our evaluation.

Following the global RMSE evaluation, we selected the top-performing model from each architecture family for detailed comparison. Table 2 presents the results of four regression metrics at 30- and 60-minute prediction horizons, following standard evaluation settings in glucose prediction tasks [15]. Specifically, we report RMSE, mean absolute error (MAE), mean absolute relative difference (MARD), and glucose-specific RMSE (gRMSE). Lower values across all metrics indicate better predictive performance. Among the evaluated LLMs, we selected the LLaMA 3.2 1B model as the core architecture of GluLLM for all subsequent experiments due to its strong performance and efficient resource usage. It is worth noting that GluLLM achieved the best performance at both prediction horizons across all regression metrics when compared with the top models from each deep learning family. Furthermore, when evaluated on the external Móstoles dataset [54], GluLLM consistently ranked first across all metrics and both time horizons, demonstrating strong generalizability and robustness to different populations and clinical contexts.

**Table 2:** Regression metrics for glucose level prediction performance (Mean*±*STD) on hold-out REPLACE-BG testing set and external Móstoles dataset at 30-minute and 60-minute prediction horizons.

|  | Method | 30 minutes |  |  |  | 60 minutes |  |  |  |
| --- | --- | --- | --- | --- | --- | --- | --- | --- | --- |
|  |  | RMSE | MAE | MARD (%) | gRMSE | RMSE | MAE | MARD (%) | gRMSE |
| REPLACE | GluLLM | **20.6 ± 3.5** | **14.7 ± 2.6** | **10.6 ± 2.4** | **25.6 ± 4.8** | **35.8 ± 6.0** | **26.6 ± 4.5** | **19.6 ± 4.5** | **46.5 ± 8.5** |
| | Crossformer | 21.1 ± 3.6 | 15.0 ± 2.6 | 10.6 ± 2.4 | 26.2 ± 4.9 | 36.3 ± 6.0 | 26.9 ± 4.5 | 19.6 ± 4.4 | 47.2 ± 8.5 |
| | NBeats | 21.4 ± 3.7 | 15.3 ± 2.7 | 10.9 ± 2.5 | 26.6 ± 5.0 | 36.6 ± 6.1 | 27.3 ± 4.6 | 19.9 ± 4.5 | 47.6 ± 8.6 |
| | TCN | 21.3 ± 3.7 | 15.2 ± 2.7 | 10.8 ± 2.4 | 26.7 ± 5.0 | 36.4 ± 6.1 | 27.2 ± 4.5 | 19.7 ± 4.4 | 47.7 ± 8.6 |
| | GRU | 22.1 ± 3.7 | 15.6 ± 2.6 | 10.9 ± 2.3 | 26.9 ± 4.9 | 38.1 ± 6.3 | 28.0 ± 4.7 | 19.8 ± 4.0 | 48.4 ± 8.8 |
| Móstoles | GluLLM | **9.6 ± 2.9** | **6.9 ± 2.1** | **6.6 ± 1.9** | **10.3 ± 3.3** | **16.0 ± 5.5** | **11.7 ± 4.0** | **11.0 ± 3.3** | **17.6 ± 6.7** |
| | Crossformer | 10.1 ± 2.9 | 7.5 ± 2.0 | 7.4 ± 2.0 | 11.0 ± 3.4 | 17.9 ± 4.6 | 14.0 ± 3.4 | 14.0 ± 3.8 | 20.4 ± 6.2 |
| | NBeats | 10.1 ± 3.0 | 7.4 ± 2.2 | 7.2 ± 2.1 | 11.1 ± 3.5 | 18.1 ± 4.7 | 14.1 ± 3.5 | 14.2 ± 4.1 | 20.7 ± 6.4 |
| | TCN | 10.2 ± 3.0 | 7.6 ± 2.1 | 7.4 ± 2.2 | 11.2 ± 3.5 | 18.5 ± 4.6 | 14.7 ± 3.4 | 14.9 ± 4.3 | 21.3 ± 6.5 |
| | GRU | 10.8 ± 3.2 | 8.1 ± 2.3 | 8.0 ± 2.5 | 12.1 ± 4.0 | 18.8 ± 4.1 | 15.3 ± 3.0 | 15.7 ± 3.9 | 21.7 ± 5.9 |

**Table 3:** Hypoglycemic event prediction performance on hold-out REPLACE-BG testing set and external Móstoles dataset.

|  | Method | ACC | PREC | SEN | SPEC | AUC | F1 | MCC |
| --- | --- | --- | --- | --- | --- | --- | --- | --- |
| REPLACE | GluLLM | **95.1** | 85.5 | **59.6** | 98.9 | **0.793** | **0.703** | **0.690** |
|  | Crossformer | 94.9 | 91.1 | 53.6 | 99.4 | 0.765 | 0.675 | 0.676 |
|  | NBeats | 95.0 | 92.1 | 53.4 | 99.5 | 0.765 | 0.676 | 0.680 |
|  | TCN | 94.8 | **94.3** | 50.4 | **99.7** | 0.751 | 0.657 | 0.669 |
|  | GRU | 95.0 | 93.1 | 52.4 | 99.6 | 0.760 | 0.671 | 0.677 |
| Móstoles | GluLLM | **97.9** | 85.4 | **68.9** | 99.4 | **0.841** | **0.763** | **0.756** |
|  | Crossformer | 97.9 | 96.9 | 59.0 | **99.9** | 0.794 | 0.733 | 0.747 |
|  | NBeats | 97.9 | 93.8 | 62.3 | 99.8 | 0.811 | 0.749 | 0.755 |
|  | TCN | 97.7 | **99.3** | 54.7 | 99.7 | 0.773 | 0.705 | 0.728 |
|  | GRU | 97.6 | 77.4 | 62.5 | 99.4 | 0.766 | 0.709 | 0.720 |

Further evaluation was conducted to assess whether the glucose predictions could effectively indicate adverse glycemic events, particularly hypoglycemia, which can lead to severe complications in a short period of time. In this analysis, we used classification metrics to determine whether the predicted glucose time series could accurately detect hypoglycemic events, defined as glucose levels below 70 mg/dL Model performance was evaluated using standard classification metrics: accuracy (ACC), precision (PREC), sensitivity (SEN), and specificity (SPEC). In addition, we included three balanced metrics commonly used in imbalanced classification tasks: area under the receiver operating characteristic curve (AUC), F1 score, and Matthews correlation coefficient (MCC)—to account for the relatively low prevalence of hypoglycemia (Supplementary Table 4). For these metrics, higher values uniformly indicate better predictive performance. We chose to report overall performance across all testing data rather than individual-level results because some participants experienced no hypoglycemic events during the monitoring period, making certain classification metrics incalculable for these individuals.

**Table 4:** Demographic characteristics (Mean *±* SD) of the individuals in the clinical datasets.

| Demographic | REPLACE-BG | Móstoles |
| --- | --- | --- |
| Age (years) | 44 ± 14 | 60 ± 10 |
| Sex (male/female) | 114/112 | 103/105 |
| Mean glucose level (mg/dL) | 159.9 ± 26.4 | 102.3 ± 12.2 |
| GMI (%) | 7.1 ± 0.6 | 5.8 ± 0.3 |
| TBR (<54 mg/dL) (%) | 1.5 ± 2.3 | 0.6 ± 2.4 |
| TBR (54-69 mg/dL) (%) | 3.5 ± 3.3 | 3.2 ± 6.2 |
| TIR (70-180 mg/dL) (%) | 62.5 ± 16.0 | 95.6 ± 7.8 |
| TAR (181-250 mg/dL) (%) | 22.5 ± 10.1 | 0.6 ± 2.4 |
| TAR (>250 mg/dL) (%) | 10.1 ± 9.9 | 0.0 ± 0.1 |
| Low blood glucose index | 1.3 ± 1.1 | 1.7 ± 1.6 |
| High blood glucose index | 7.6 ± 4.4 | 0.3 ± 0.5 |
| Inter-day CV (%) | 37.1 ± 8.6 | 16.7 ± 5.5 |
| Intra-day CV (%) | 34.0 ± 7.8 | 16.7 ± 5.5 |

GMI: glucose management indicator, TBR: time below range, TIR: time in range, TAR: time above range, CV: coefficient of variation

GluLLM achieved the highest scores for AUC, F1, and MCC, indicating the best overall classification performance among all models. Notably, GluLLM also significantly improved sensitivity, detecting a greater proportion of hypoglycemic events on both datasets: an improvement of 11.2% on the REPLACE-BG dataset and 10.2% on the Móstoles dataset compared with the second-best models, Crossformer and GRU, respectively. These gains suggest that implementing GluLLM could potentially lead to a significant reduction in the occurrence of hypoglycemic events.

### Memory and CPU footprint of GluLLM on smartphone platforms

Smartphone platforms are widely used in modern glucose management systems, typically paired with CGMs via Bluetooth. However, implementing LLMs on smartphones presents significant challenges due to the substantial size of these models and the quadratic computational complexity of Transformer blocks. Despite these constraints, on-device and offline LLMs are essential in healthcare applications, where data privacy and continuous availability are critical.

To demonstrate feasibility, we implemented GluLLM on two smartphone devices: the iPhone 14 Pro Max (representing a mainstream high-end device) and the iPhone SE 2020 (representing a resource-constrained platform). Fig. 3 shows the peak CPU and memory usage during GluLLM inference on both devices. Peak values are reported because sudden computational spikes can exceed device capabilities and risk freezing or crashing the system. The iPhone 14 Pro Max consumed a maximum of 55.5% CPU and 453.9 MB of memory (7.4% of the device’s total 6GB memory capacity), while the iPhone SE 2020 consumed a maximum of 85.8% CPU and 512.2 MB of memory (16.7% of the device’s total 3GB memory capacity). When examining the computational demands of individual components, we found that the encoder and decoder modules required substantially fewer resources compared with the time dependent interpreter (TDI) module and LLM body, consuming a maximum of 24.7% CPU and 10.2 MB of memory on the iPhone 14 Pro Max, and 36.6% CPU and 19.7 MB of memory on the iPhone SE 2020.

**Fig. 3:**
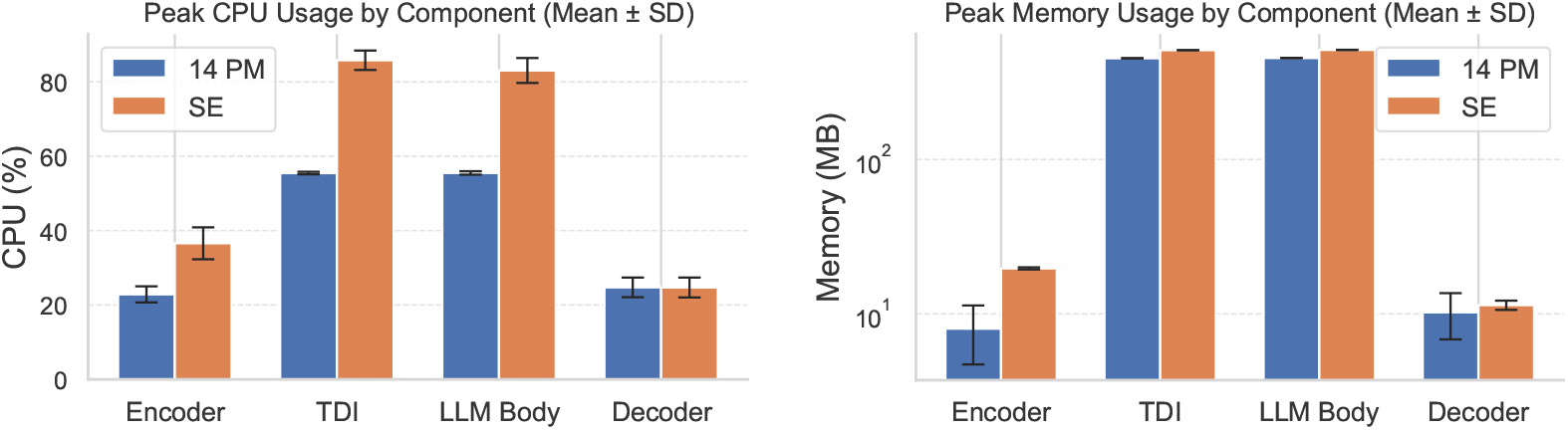
Peak CPU and memory usage of GluLLM across smartphone platforms. Blue and orange bars represent the performance of the iPhone 14 Pro Max (14PM) and iPhone SE 2020 (SE), respectively. Error bars indicate the mean *±* standard deviation across 10 independent runs.

The average memory consumption for running GluLLM was 139.1 MB (2.5%) on the iPhone 14 Pro Max and 178.1 MB (5.8%) on the iPhone SE, both exhibiting low energy impact. All operations were completed within 20 and 35 seconds on the 14 Pro Max and SE, respectively. These results demonstrate that deploying GluLLM on smartphone platforms for real-time clinical decision support is feasible.

## Discussion

In this work, we proposed GluLLM, an adapter framework that enhances pretrained LLMs for digital health management, with a specific focus on glucose prediction. To our knowledge, this is the first study to introduce an on-device LLM framework that integrates multimodal data, including wearables, daily event logs, and electronic health records (EHRs), to support comprehensive and personalized care. We conducted a thorough evaluation by benchmarking against 15 state-of-the-art deep learning approaches for time series forecasting. The proposed GluLLM with LlaMA 3.2 1B model achieved the best regression performance and superior results in hypoglycemia detection, significantly increasing sensitivity to these adverse events. Further implementation on smartphone platforms demonstrated the feasibility of real-world deployment to enhance clinical decision support systems with minimal resource requirements.

The performance gains achieved by GluLLM can be attributed to several key design elements. First, incorporating contextual information significantly enhances LLM performance in time series forecasting, as shown in prior work such as TimeLLM [26]. In our case, demographic characteristics closely related to an individual’s metabolic status, such as age, sex, BMI, and HbA1c, substantially influence glucose dynamics. Integrating this information enables the personalization of a population-level model, allowing it to generalize effectively to unseen individuals without the need for fine-tuning or post-training, as demonstrated in our ablation study on the Móstoles dataset (Supplementary Fig. 6). Second, the autoregressive architecture and next-token prediction framework allow GluLLM to translate token transitions learned during pretraining into meaningful glucose trajectories over time, which leverages the core strengths of language models. Finally, the TDI module enables real-time adaptation by integrating textual inputs describing daily activities. This allows the model to account for transient glucose fluctuations, such as those resulting from insulin interventions, thereby improving predictive accuracy (Supplementary Fig. 6). With this setup, GluLLM enables largescale deployment of personalized models across diverse population cohorts, without the need for individual-specific training. It can be readily integrated into a variety of clinical settings to deliver robust and generalizable performance. Moreover, it is straightforward to incorporate data from multiple wearable sensors through additional encoder modules, such as combining wristband sensors with CGMs [19], to enable more comprehensive physiological monitoring and further enhance predictive accuracy. The proposed framework is also extensible to other health condition management tasks, as well as biomarker and vital signs monitoring. For example, it could be applied to wearable-enabled Parkinson’s disease care [55] by incorporating data from wearable accelerometers and EHRs to support holistic and personalized care.

Although LLM-driven approaches may appear computationally intensive for traditional forecasting tasks, they are particularly well suited to biomedical time series, which are often accompanied by rich textual information that directly influences prediction. This synergy between structured signals, such as wearable data, and unstructured context, such as EHR notes, makes LLMs especially powerful. These capabilities are further enhanced by the highly efficient design of GluLLM. The training complexity is substantially lower than that of full large language model fine-tuning or even many parameter-efficient fine-tuning (PEFT) approaches [56], such as LoRA [57]. We chose to use neural network–based adapters rather than PEFT methods, because the primary goal of our task is to improve how the model interprets input data from wearable devices, rather than to enhance the generation of natural language responses. In our context, the quality of the input representation is more critical than producing fluent textual outputs. Future work will explore integrating LoRA into GluLLM to investigate whether further improvements can be obtained. In particular, if predicted values are to be used for clinical decision support, such as suggesting diagnoses or feasible treatment plans based on model predictions, LoRA will be employed to fine-tune response generation. Such text outputs could then be evaluated in collaboration with clinicians to assess their utility in real-world healthcare settings. Moreover, generating explanatory text alongside time series predictions may provide reasoning and insight into why the model produces certain forecasts [58], representing an important direction for our future investigation in model interpretability.

Beyond training efficiency, we also demonstrated the feasibility of deploying GluLLM in resource-constrained smartphones. This is particularly important for preserving the privacy of sensitive health data, as it ensures that personal information remains on the user’s own device. On-device deployment is a particularly viable approach for LLMs in healthcare applications that require continuous, high-demand management tasks. In addition, clinical prediction and decision support are inherently time sensitive, and people cannot afford service interruptions, especially in scenarios where Internet connectivity is limited or unstable. This limitation is a common issue in cloud-based LLM deployments, further highlighting the importance of enabling robust, offline-capable inference on edge devices. As shown in Fig. 3, LLM body and the TDI module are the primary contributors to CPU, memory, and power consumption during model execution. However, it is important to note that if the contextual information processed by TDI were instead fed directly into the LLM, the significantly longer input sequence would substantially increase computational demands. This is due to the quadratic complexity of Transformer layers with respect to input length. By separating daily contextual information into the TDI module, we not only preserve modeling flexibility but also improve both training and inference efficiency. Furthermore, in many cases, particularly when no insulin has been administered in the last hour, the embedding of TDI remains unchanged over days. In such scenarios, the embedding can be precomputed and stored on the device, eliminating the need for repeated inference and further reducing computational overhead. The total consumption of GluLLM resources on a smartphone is feasible, demonstrating its potential for long-term clinical decision support on devices.

Moreover, the lightweight nature of the encoder and decoder modules, consuming only minimal system resources, makes it practical to deploy multiple neural network-based adapters for managing various health conditions in a unified digital health system (Fig. 1). A further step toward advancing model deployment is to perform parameter quantization [59], which compresses the model and accelerates inference by reducing storage and computational demands. This approach would enable LLM-driven health management to be implemented on devices with more resources constraints and could also allow the use of models with larger parameter sizes for more powerful capability under limited hardware conditions.

## Methods

### Multimodal data preprocessing

In this work, we used two primary clinical datasets with varied patient conditions. The first is REPLACE-BG [52], containing data from 226 participants with T1D at 14 US sites. Participants were equipped with Dexcom G4 CGM during a 26-week trial period. The second dataset was collected by University Hospital of Móstoles [54] in Madrid, comprising data from 207 participants with previous diagnoses of essential hypertension and explicit exclusion of prior diabetes diagnoses or antidiabetic medication use. These participants were monitored using Medtronic MiniMed iPro devices for periods ranging from 24 hours to 3 days, with a 33-month follow-up identifying 17 new cases of T2D. Both datasets are publicly available and provide comprehensive demographic information on participants, with glucose readings sampled at five-minute intervals. In particular, REPLACE-BG includes insulin bolus data, which is absent from the Móstoles dataset, since insulin treatment was not used for participants without diabetes. Supplementary Table 4 summarizes the demographics of these datasets. It is worth highlighting the significant differences between them, particularly in mean glucose levels and time in target range (TIR), underscoring the importance of model generalization capabilities for effective cross-dataset performance.

To provide comprehensive prediction, we utilized all available information from this complex health management scenario. This includes CGM data as a time series, self-reported or device-logged daily events, and EHRs. CGM data was first scaled using standardization. Although both datasets maintain high quality with minimal missing CGM readings, we apply a mixed interpolation and extrapolation strategy, as used in our previous work [21]. Specifically, interpolation is applied when missing values occur within the middle of an input sequence, while extrapolation is used when gaps appear at the end, thereby avoiding data leakage. Demographic information was extracted and formatted for input to the EHR prompt module as: “Characteristics: - Age: {age}, - Sex: {sex}, - BMI: {bmi}, - HbA1c: {hba1c}, Predict next value based on historical glucose embedding:”. Timestamp data from CGM was formatted in date format and combined with insulin bolus information (when available) as input to the TDI module, for instance: “Historical data from 13:30 to 14:30. 5 units of bolus insulin delivered.” In the context of glucose prediction, we utilize historical CGM data within a defined temporal window to forecast future glycemic trajectories. Specifically, at any timestep *t*, we process a sequence of historical glucose readings **G**_*t*_ = [*g*_*t*−*L*+1_, *g*_*t*−*L*+2_, …, *g*_*t*_] ∈ ℝ^*L*^, where *L* represents the fixed observation window. This historical data, combined with relevant clinical and contextual information, serves as input to predict the corresponding future glucose sequence **Ĝ** _*t*_ = [*ĝ*_*t*+1_, *ĝ*_*t*+2_, …, *ĝ*_*t*+*τ*_ ] ∈ ℝ^*τ*^ where *τ* denotes the prediction horizon. This mathematical formulation establishes the foundation for our sequence-to-sequence modeling approach.

### GluLLM for multimodal data integration

We first utilized the tokenizer and embedding modules from the original LLM architecture to convert the EHR prompt into embedding representations, following standard natural language processing procedures. The parameters of the LLMs are frozen and no training is needed at this stage. We denote this embedding output as **EE**. It is important to note that demographic information is considered consistent for each individual in our experiments, allowing this embedding operation to be performed just once per individual, which improves computational efficiency. This prefix-prompt approach was motivated by recent advances in prompt engineering that have demonstrated significant improvements in LLM generalization performance, particularly for medical applications that may not have been encountered during model training [60]. This technique also represents a core module in TimeLLM [26], which has shown a substantial reduction in error for time series forecasting across various scenarios.

Subsequently, we converted historical glucose readings **G**_*t*_ into representations similar to language embeddings to enable compatibility with the LLM architecture. Various methods exist in the literature for this dimension alignment, ranging from simple linear layers [61] to more complex multi-head attention mechanisms [26]. Inspired by a recent efficient approach proposed in AutoTimes [27], which effectively captures rich chronological information, we divided historical CGM data into segments, each represented as a token. These tokens were then converted by the GluLLM encoder module to embeddings with dimensions that match the embedding vectors of the language tokens. We implemented fully connected neural networks to map this dimensional relationship. Although the length of the segments could be arbitrary, we used the prediction length *τ* as the segment length to naturally reformulate the problem into next-token prediction, which is a standard training objective for decoder-only LLMs [34, 39, 62]. This approach allowed us to leverage the powerful predictive capabilities and token transition inherent in these language models. Additionally, we assumed that the length of historical CGM data *L* was *N* multiples of *τ* to ensure consistent and uniform segmentation throughout the analysis. In this case, the input and output in glucose prediction can be rewritten as 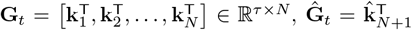, where **k**_*n*_ = *g*_*t*+(*n*−*N* −1)*τ* +1_, *g*_*t*+(*n*−*N* −1)*τ* +2_, *g*_*t*+(*n*−*N* −1)*τ* +*τ*_. Here, we set *L* = 72 and *τ* = 12, corresponding to six hours of historical CGM readings and a one-hour prediction horizon. The six-hour window captures the typical duration of insulin action, as the effects of a meal insulin bolus generally subside within this period. A one-hour prediction horizon is widely adopted in glucose prediction applications [15], which allows individuals sufficient opportunity to take proactive interventions in response to anticipated glycemic excursions.

After converting each token **k** in the GluLLM encoder, we obtain the glucose embedding;

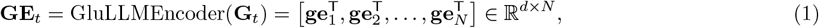

where *d* denotes the dimensionality of the LLM’s embedding, which varies across the different models considered in this work. To fully leverage temporal information, we also divided the timestamp and insulin data into the same segments used for the CGM data, and input them into the TDI. As illustrated in Fig. 1, TDI comprises a tokenizer, an embedding layer, and the LLM body from the original LLM. This design allows the model to process temporal information and associated daily activities, producing a corresponding language embedding. Specifically, we selected the last hidden state vector from the output embedding of each segment, as this position typically aggregates the most relevant contextual information from the input text and is commonly used in LLMs to predict the probability of the next token. By concatenating these vectors, we obtain 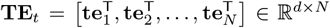. Aligning the temporal embedding vectors with the glucose embedding vectors in a one-to-one fashion, similar to positional encoding, and prepending the EHR prompt embedding at the beginning, we construct the final input embedding for the LLM. The output embedding is then obtained by processing this input through the LLM body:

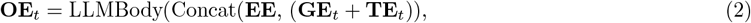

where the addition **GE**_*t*_ + **TE**_*t*_ is performed element-wise. In this regard, the next-hour of glucose levels can be obtained by projecting the last hidden hidden state using GluLLM decoder, which is also implemented as a fully connected neural network, and we have

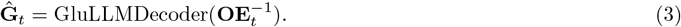

The flexibility of next-token prediction in this model also enables us to extend the prediction horizon beyond one hour. For example, to achieve a two-hour prediction horizon, we simply append the previously predicted glucose token to the input and perform an additional round of next-token prediction. Preliminary results, shown in Supplementary Fig. 5, demonstrate that this approach achieves over 90% clinically acceptable predictions at the extended two-hour horizon in both datasets.

### Model training and evaluation

To train a population-level glucose prediction model, we partitioned the data on a per-individual basis to ensure subject-level separation. Given the substantial volume of data in the REPLACE-BG dataset (approximately 12 million CGM readings), we used it for model training, validation, and hold-out testing, while the Móstoles dataset served as an external validation cohort. Specifically, the REPLACE-BG dataset was first divided into a development set comprising 180 individuals and a hold-out testing set of 46 individuals. We then performed five-fold cross-validation on the development set to fine-tune the hyperparameters of GluLLM. In each fold, data from 135 individuals were used for training, while the remaining 45 individuals were used for validation. This process was repeated across folds to ensure robust hyperparameter selection (Supplementary Fig. 4).

The performance of the model was evaluated primarily using the global RMSE over the prediction horizon *τ*. In the detailed comparison, RMSE is the primary metric reflecting overall prediction error at a specific time point. MAE is also widely used and is known to exhibit strong discriminative ability for glycemic variability [63]. MARD is a well-established accuracy metric in the CGM industry [64]. Finally, gRMSE is a customized RMSE that incorporates a penalty function inspired by the Clarke Error Grid [65], emphasizing clinical relevance by penalizing the prediction errors that could lead to harmful therapeutic interventions. The detailed definitions and equations for these metrics are provided in Supplementary Information.

All deep learning models were implemented in PyTorch and accelerated using NVIDIA A10 Tensor Core GPUs. Hyperparameter optimization was conducted using Optuna to ensure efficient and systematic tuning. We performed a maximum of 10 optimization trials per model, exploring a search space that included learning rate, batch size, hidden layer dimensions, dropout rate, and optimizer parameters. After identifying optimal hyperparameters, we trained the model on the entire development set and evaluated its performance on the independent hold-out testing set of 46 individuals and the external validation cohort from the Móstoles dataset, which includes 207 individuals. Notably, hyperparameter tuning was performed for all baseline models to ensure fair comparisons. We employed the Adam optimizer and the mean square error loss function to optimize the model parameters.

The trainable modules in GluLLM consist only of the encoder and decoder components, which comprise stacks of fully connected neural network layers. These components have a significantly smaller number of trainable parameters compared to full LLM fine-tuning or even PEFT approaches [56]. The computational complexity of this design is comparable to that of an extreme case of LoRA-based adapters [57], where a low-rank matrix with rank 1 is applied to a single Transformer layer. However, even with this efficient design, training GluLLM with larger models such as LLaMA 8B and Mistral 7B remains challenging on a GPU with 24 GB of RAM, even with a single batch of data, due to the significant computational demands of LLM inference. To address this, we distributed the Transformer layers of these two models across 8 NVIDIA A10 Tensor Core GPUs, the maximum number of GPUs available on a single node of our high-performance computing cluster, to enable model training. For smaller LLMs, inference can be performed on a single GPU; thus, we employed standard data parallelism by distributing batches across multiple GPUs to accelerate training.

### Smartphone platform implementation

One of the most challenging aspects of deploying LLMs for on-device health management is determining the feasibility of implementation. To address this concern, we conducted a study implementing GluLLM on smartphone platforms. We selected the iOS platform specifically, as on-device LLMs will be available in their next generation of iPhones. For development, we utilized Swift 5 and Xcode 16.2 as the integrated development environment. It should be noted that we focused solely on evaluating memory and CPU usage of the proposed LLM-driven health application rather than developing a complete user interface.

We implemented LLaMA 3.2 1B model using the GGUF format from Huggingface [66] with Full F16 weights and without quantization. The LLM inference was handled using the llama.cpp library [67]. For integration, the GluLLM encoder and decoder components were converted to Core ML format to facilitate efficient interaction with the LLaMA model. Real-time CPU and memory usage were assessed using Xcode’s profiling tools. The evaluation was conducted on an iPhone 14 Pro Max, equipped with an Apple A16 Bionic chip, a 6-core CPU, and 6 GB of RAM, as well as on an iPhone SE (2020), featuring an A13 Bionic chip, a 6-core CPU, and 3 GB of RAM.

## Data availability

Datasets used in this study are publicly available. The REPLACE-BG dataset was obtained from Aleppo *et al*. (2017), “REPLACE-BG RCT Protocol 10-27-15 v2.0,” available at https://public.jaeb.org/dataset/546. The analyses, interpretations, and conclusions presented herein are solely the responsibility of the authors and have not been reviewed or endorsed by Aleppo *et al*. The Móstoles dataset was obtained from Colás *et al*. (2019), available at https://journals.plos.org/plosone/article?id=10.1371/journal.pone.0225817.

## Code availability

This study was implemented using the free and open-source programming language Python (version 3.11.8) along with the PyTorch (version 2.1.2) deep learning framework. The source code for both the deep learning model and the smartphone app is available from the corresponding authors upon reasonable request.

## Acknowledgements

Taiyu Zhu is fully funded by Novo Nordisk and is a Novo Nordisk Postdoctoral Fellowship run in partnership with the University of Oxford.

## Supplementary

## Participant demographic profiles in the two clinical datasets

Table 4 presents the demographic characteristics of participants in the REPLACE-BG and Móstoles datasets.

## Cohort stratification and cross-validation framework

Fig. 4 illustrates the cohort stratification used in the five-fold cross-validation framework, as well as the procedures for model training and hold-out testing.

**Fig. 4:**
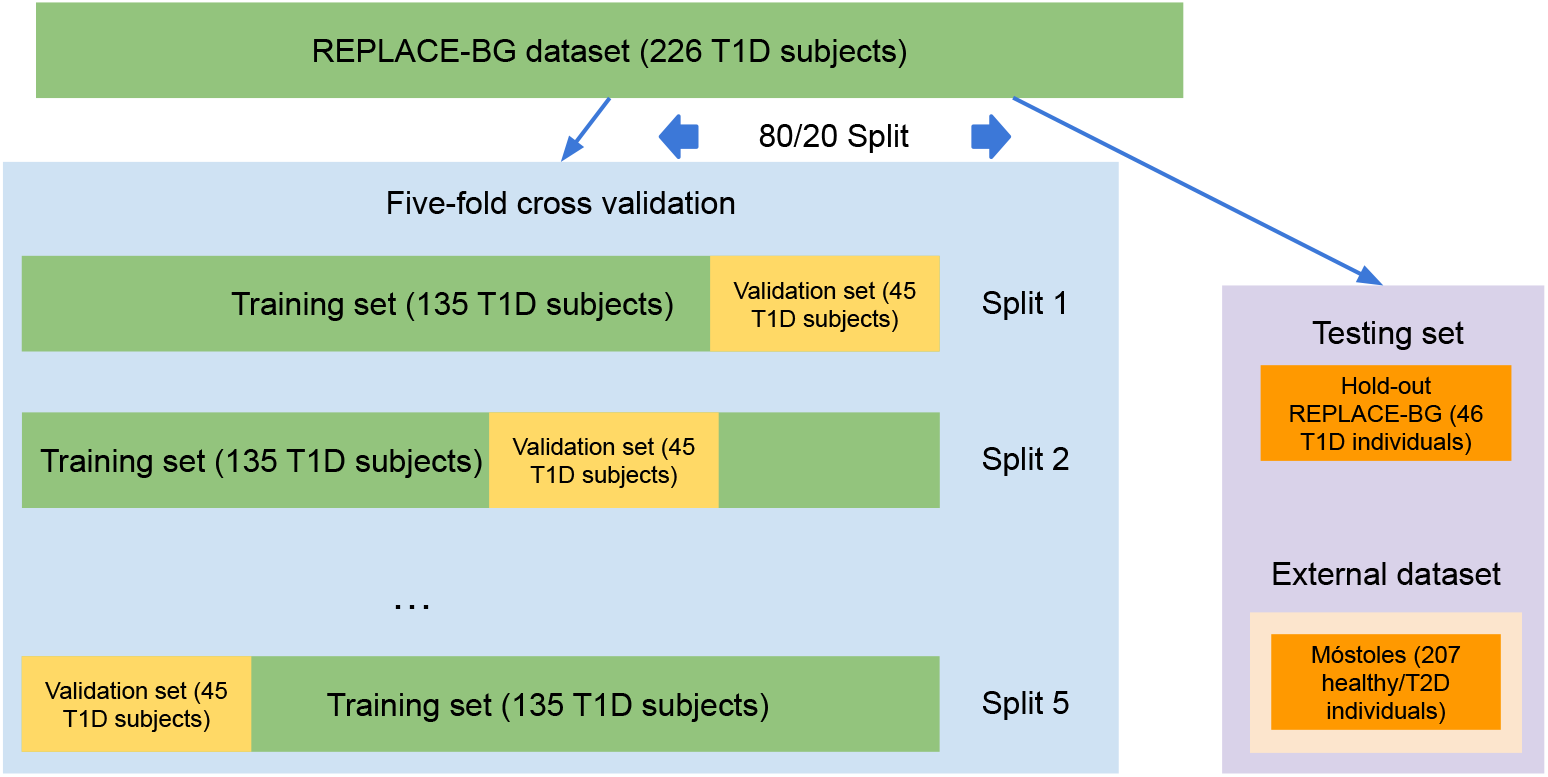
Data partitioning strategy. The REPLACE-BG dataset was first divided into a development set, which was used for training and validation in five-fold cross-validation, and a hold-out testing set, with splitting performed on an individual basis to prevent data leakage. The Móstoles dataset served as an independent external validation cohort to assess generalization performance.

## Definition of regression metrics

Three widely used regression metrics for glucoselevel prediction were empolyed: RMSE, MAE, and MARD, defined as:

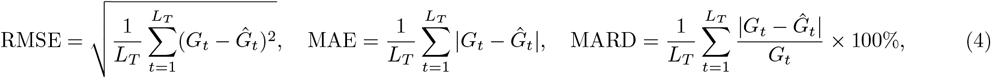

where *L*_*T*_ denotes the total number of testing samples, *G*_*t*_ is the ground truth BG measurement at time *t*, and *Ĝ*_*t*_ is the corresponding predicted value.

To assess clinical relevance, we further used gRMSE [65] as follows:

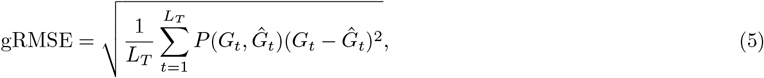

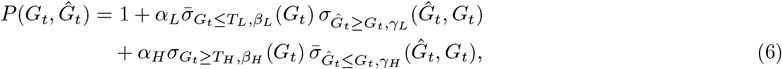

where *P* is a penalty function inspired by the Clarke Error Grid (CEG) [68], designed to penalize overestimation in hypoglycemia and underestimation in hyperglycemia. Definitions of the sigmoid-like functions 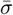, *σ*, and the corresponding parameters [*α*_*L*_, *β*_*L*_, *γ*_*L*_, *T*_*L*_, *α*_*H*_, *β*_*H*_, *γ*_*H*_, *T*_*H*_ ] can be found in the literature [65].

## Clinically-relevant analysis at extended two-hour prediction horizon

The percentage of predictions falling within zones A+B, which indicate clinically acceptable accuracy, was very high across both datasets. For the REPLACE-BG cohort, GluLLM achieved 92.26% accuracy within these zones, while for the Móstoles dataset, it reached an exceptional 99.09% accuracy as shown in Fig. 5. These results demonstrate the strong clinical utility of our predictions even at extended forecast horizons. We did not provide a CEG plot for the REPLACE-BG dataset due to the excessive density of data points, which would render the visualization difficult to interpret.

**Fig. 5:**
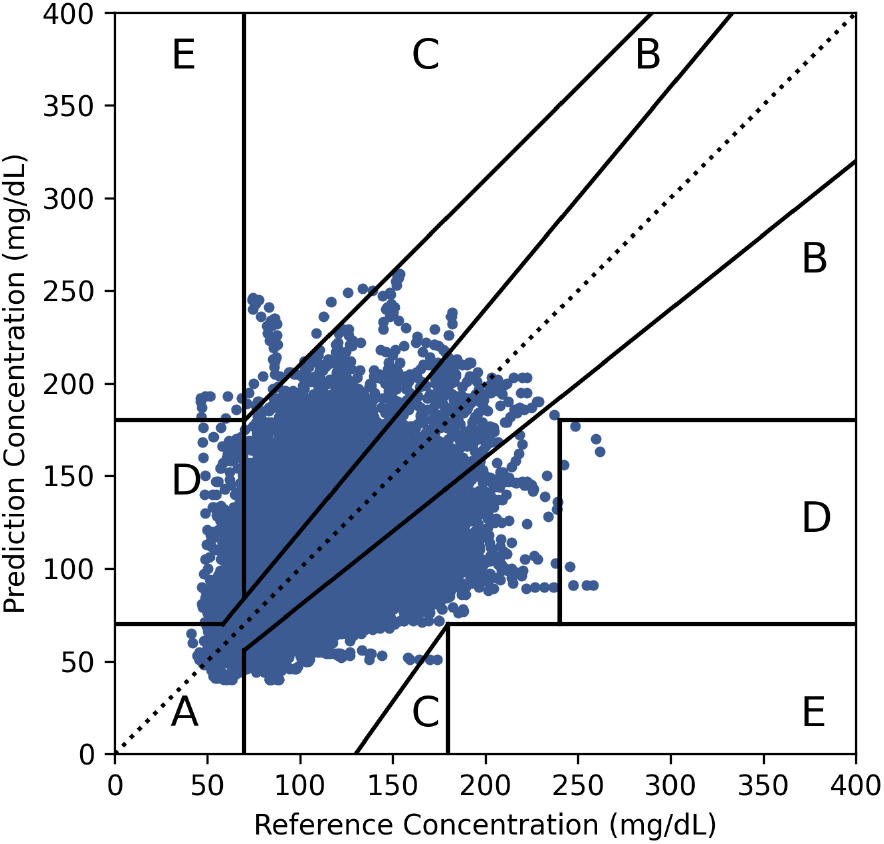
Clark error grid (CEG) analysis comparing predicted glucose values against actual measurements at two-hour prediction horizon. This clinically-relevant visualization categorizes prediction errors into zones reflecting varying degrees of clinical significance, with zones A and B representing clinically acceptable predictions and zones C-E indicating potentially dangerous errors that could lead to inappropriate treatment decisions.

## Ablation study

We conducted an ablation study to investigate the contribution of each central module in GluLLM, as shown in Fig. 6. The results show that each module effectively reduces the global RMSE across both datasets. Notably, the improvement contributed by the TDI module is smaller in the Móstoles dataset compared to the REPLACE-BG dataset. This is primarily because the TDI input in REPLACE-BG includes insulin information, reflecting the intensive management for individuals with T1D, whereas such information is not available in the Móstoles dataset.

**Fig. 6:**
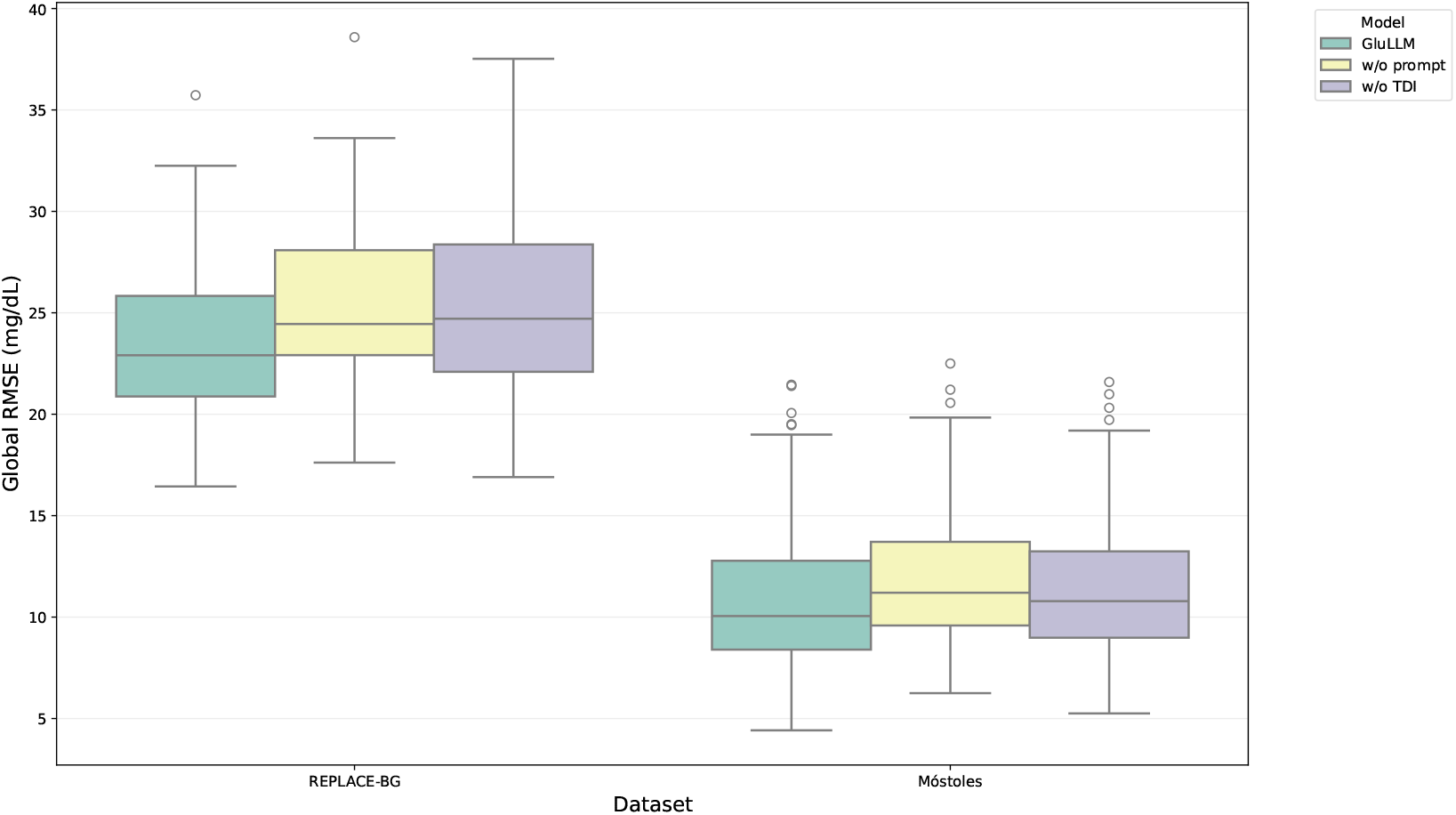
Ablation study evaluating the contributions of the EHR prompt and TDI modules to glucose prediction performance. Global RMSE over a 60-minute prediction horizon is used as the evaluation metric. Each dot represents the RMSE for an individual participant in the respective dataset. In the box plots, the lower and upper hinges correspond to the first (Q1) and third (Q3) quartiles, respectively; the central line indicates the median; and the whiskers extend to 1.5 times the interquartile range (IQR). Circles denote statistical outliers.

